# A Mendelian randomization study with clinical follow-up links metabolites to risk and severity of pulmonary arterial hypertension

**DOI:** 10.1101/2023.06.30.23292100

**Authors:** Elham Alhathli, Thomas Julian, Zain Ul Abideen Girach, A A Roger Thompson, Christopher Rhodes, Stefan Gräf, Niamh Errington, Martin R Wilkins, Allan Lawrie, Dennis Wang, Johnathan Cooper-Knock

## Abstract

Pulmonary arterial hypertension (PAH) exhibits phenotypic heterogeneity and variable response to therapy. The metabolome has been implicated in the pathogenesis of PAH, but previous works have lacked power to implicate specific metabolites. Mendelian randomisation (MR) is a method for causal inference between exposures and outcomes. Using GWAS summary statistics, we implemented hypothesis-free MR methodology to test for causal relationships between serum concentration of 575 metabolites and PAH. Unbiased MR causally associated five metabolites with risk of PAH after stringent multiple testing correction; of the five candidates, serine and homostachydrine were validated in a different larger PAH GWAS, and associated with clinical severity of PAH via direct measurement in an independent clinical cohort of 449 PAH patients. We used conditional and orthogonal approaches to explore the biology underlying our lead metabolites. A rare variant analysis demonstrated that loss of function (LOF) mutations within ATF4, a transcription factor responsible for upregulation of serine synthesis under conditions of serine starvation, are associated with higher risk for PAH. Homostachydrine is a xenobiotic metabolite that is structurally related to L-proline betaine, which has been previously linked to modulation of inflammation and tissue remodelling in PAH. Our MVMR analysis suggests that the effect of L-proline betaine is actually mediated indirectly via homostachydrine. Our data presents a new method for study of the metabolome in the context of PAH, and suggests several candidates for further evaluation and translational research.

## Introduction

Pulmonary arterial hypertension (PAH), formerly known as primary pulmonary hypertension, is a rare life-threatening disorder of the pulmonary circulation. It is an archetypal complex disease caused by the interaction of risk genotypes with the environment^1^. Significant advances have been made to understand genetic and environmental risk factors for PAH^2^, but much is still unknown, which limits efforts to develop effective treatments. Certain genetic susceptibility genes have been identified but it is currently not possible to predict who will be affected or when the disease will occur. Mendelian randomisation (MR) is a method for studying the effects of exposures on specific outcomes, such as disease risk, using genetic variation within a population as a natural experiment^3^. MR is an example of a causal inference methodology, whereby a significant relationship implies a causal effect of the exposure of interest on the disease trait, providing a series of assumptions are met^3^. Genetic variation is fixed at conception and therefore SNPs, which form the instruments used in MR, can be considered upstream of adult-onset disease^4^ such as PAH.

The metabolome is an integrator of genetic and environmental factors; it reflects the state of tissues and cells and can alter their function. As such the metabolome is a good candidate to explore the biological mechanisms underpinning PAH, and indeed there is evidence for the role of the metabolome in the risk and severity of PAH. For example, Rhodes, Ghataorhe, *et al.* (2017) examined 686 biological metabolites in 365 individuals with idiopathic/heritable PAH, 121 healthy controls, and 139 disease controls using ultraperformance liquid chromatography mass spectrometry^5^. Sets of 20-50 metabolites distinguished patients with PAH from controls and disease controls, and correlated with disease severity as measured by survival. Expanding this investigation to patients with chronic thromboembolic PH demonstrated comparable metabolic abnormalities despite their different underlying biological dysfunction; interestingly these patients also experienced considerable restoration of their metabolic profiles after therapeutic pulmonary thromboendarterectomy surgery^6^.

To date studies of the metabolome in PAH described have suffered from limited power because of relatively small sample sizes; as an example, the study described above highlighted a panel of metabolites^5^ but was underpowered to implicate individual metabolites in PAH pathogenesis. This is largely a result of a requirement for direct measurement of metabolite concentrations in serum from a large number of PAH patients. By using MR we overcome this key bottleneck by using genetic correlates to infer metabolite concentrations within PAH patients, thereby negating the requirement for direct measurement. By utilising different populations to evaluate exposure and outcome, so-called ‘two-sample’ MR, can achieve large sample sizes without the need to perform all measurements in the same sample set^3^. In the same way, MR also alleviates any bias in the selection of subjects which often hampers observational studies^3^. Extensive GWAS data are available to reveal the genetic architecture of the metabolome^7,8^; variants identified in this way can be used as instruments for MR. We have previously used MR to perform an hypothesis-free search for metabolites and here we apply the same methodology to PAH; we identify five metabolites which are linked to PAH after stringent multiple testing and we provide independent support for the effect of two of these metabolites on the clinical severity of PAH **(Figure 1)**. Our work opens important new avenues in the search for an understanding of the biological mechanisms underpinning PAH.

**Figure 1:**
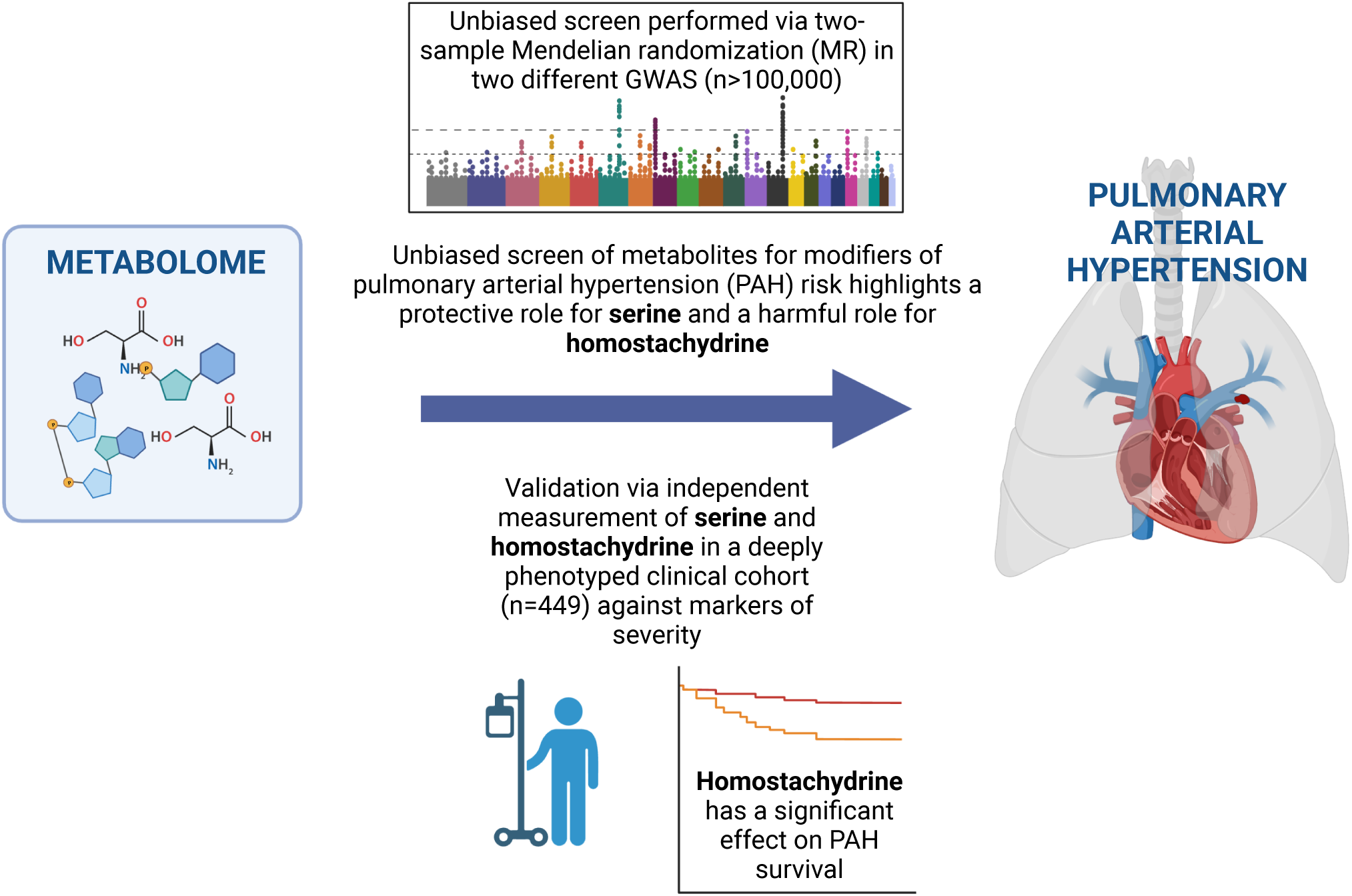
Overview of the unbiased screen for metabolites which modify risk of PAH. We performed an unbiased two-sample MR screen of 575 metabolites in order to identify those causally linked to PAH in a FinnGenn Cohort. Metabolites which passed FDR multiple testing correction were further evaluated by robust MR measures, sensitivity analysis and in a validation FinnGenn PAH GWAS. Finally, we directly measured these metabolites in an independent UK PAH cohort of 449 patients and demonstrated a correlation with disease severity.

## Methods

### Exposure and outcome GWAS

We used two-sample MR to determine whether metabolite exposures are causally linked to the risk of PAH in a FinnGen PAH GWAS of 125 cases and 162,837 controls (https://www.finngen.fi/en). A more recent FinnGen PAH GWAS of 208 cases and 243,756 controls was used to confirm metabolites which passed FDR multiple testing correction in the first GWAS^9^.

We performed a hypothesis-free analysis of the effect of 575 serum metabolites on PAH risk using publicly available GWAS studies of the serum metabolome^7,8^. The two source studies differed in sample size and methodology: Kettunen *et al* employed NMR spectroscopy to quantify metabolites in serum samples from up to 24,925 individuals^8^; whereas Shin *et al* employed liquid-phase and gas chromatography coupled with tandem mass spectroscopy in serum from 7,824 individuals^7^. The sample population for both studies was almost exclusively European; both studies included ∼2,000 individuals from the KORA cohort (Cooperative Health Research in the Region of Augsburg). The different methodologies employed were complementary: NMR is more easily applied in a high-throughput manner but sensitivity is lower than mass spectroscopy-based quantification^10^. A total of 16 metabolites (4% of the total number) were measured in both source studies not including any of our five top candidates.

We performed a second hypothesis-free analysis of the effect of 150 immune traits on PAH risk. Here we utilised a separate publicly available GWAS of blood concentration of a set of human immune system traits in 669 female twins^11^. Immune cells were quantified via specific antibodies and flow cytometry.

### Two-sample Mendelian Randomisation (MR)

Genetic instruments were selected based on an arbitrary p-value ranging from p < 5e-08 to p < 5e-06, which was varied to ensure a minimum of five instrumental SNPs. When the cut-off is too low, informative instruments will be lost, but when it is too high, non-informative instruments will be introduced and instrument pleiotropy is more likely to occur^12^. For each metabolite and immune trait studied in our unbiased screens, we performed an MR using a multiplicative random-effects inverse-variance weighted (IVW) estimate for significance testing, as this method has the greatest statistical power and it allows us to make accurate causal inferences for >4 instrumental SNPs under the assumption of limited balanced pleiotropy. IVW estimates were FDR corrected for multiple testing. Independent SNPs were clumped using a stringent cut-off of R < 0.001 within a 10,000 kb window in the European reference panel. For SNPs in LD, the SNP with the lowest p-value was retained. When an exposure SNP was unavailable in the outcome dataset, a proxy with a high degree of LD (R2 > 0.9) within a European reference population was identified. SNP effects on outcomes and exposures were harmonised so that the beta values are based on the same alleles. In order to reduce the risk of errors due to strand issues, palindromic alleles with minor allele frequency (MAF) >0.42 were excluded from the analysis^12^.

### Robust tests and sensitivity analysis

In order to increase confidence in the IVW results from our unbiased screening, we performed a series of robust MR measures and sensitivity analyses. We used an F-statistic to measure the strength of the association between instrumental SNPs and the exposure of interest. An F-statistic > 10 indicates that an SNP-derived estimate has a bias of < 10% of its intragroup variability and signifies an acceptable instrument. Pleiotropy occurs between SNPs where the difference in effect size for the exposure is not proportional to the difference in effect size for the outcome, and is usually due to a violation of one of the key assumptions underlying MR, that instrumental SNPs should be associated with the outcome only through the exposure^3^. To account for pleiotropy, we removed SNPs where the p-value for the association with the outcome was lower than for the association with the exposure of interest. The MR-Egger intercept test was also used to identify directional horizontal pleiotropy. To avoid false positives, we limited our analysis to exposures with > 10 instrumental SNPs and used a q-q plot to determine whether the median p-value was significantly inflated. A test with too few SNPs gives excessive weight to each individual SNP^13^. As IVW estimates are vulnerable to pleiotropic SNPs, we used Cochran’s Q test (p > 0.05) as a sensitivity measure to detect heterogeneity indicating pleiotropy. Moreover, radial-MR ^14^ was used to remove statistically significant outlier SNPs. The *I*^2^ statistic was used to measure the heterogeneity between variant-specific causal estimates, with a low *I*^2^ indicating bias toward the null hypothesis (Burgess and Thompson, 2017). A leave-one-out (LOO) analysis was applied to identify results where one or more SNPs exert a disproportionate effect. TwoSampleMR (version 0.5.6), Mendelian Randomization (version 0.5.1) and RadialMR (version 1.0) R packages were used for all MR analyses.

### Clinical Validation

We evaluated the identified serum metabolites in MR results from the FinnGen GWAS study using measurements of the same molecules within plasma samples from the UK PAH cohort^5,6^ including 449 PAH patients. In this cohort, metabolic profiling was performed using ultra-performance liquid chromatography mass spectrometry^5,6^. We examined the concentration of serine and homostachydrine and tested for correlation between identified candidates and clinical measures of PAH severity including survival. In all analyses we adjusted for other covariates such as age, gender, BMI, ethnicity and geographic location of sample collection.

Clinical measures of severity in the UK PAH cohort included: WHO functional class (WHO-FC); N-terminal pro-brain natriuretic peptide (NT-proBNP) which correlates with myocardial stress and provides prognostic information; six-minute walking distance (6MWD); and a series of measurements taken at right-heart catheterisation including pulmonary arterial wedge pressure (PAWP), pulmonary vascular resistance (PVR), right atrial pressure, cardiac index (CI), cardiac output (CO) and venous oxygen saturation. We also included the percentage of predicted forced vital capacity (FVC) as a measure of lung function. REVEAL is an aggregate score used for prognostication in PAH^15^. Many of these measures are not specific to PAH but have been associated with clinical outcomes including survival. In the analysis of clinical measurements we utilised linear/logistic regression adjusted for other covariates such as age, gender, BMI, ethnicity and geographic location of sample collection. In the analysis of survival we applied Cox regression, with adjusting for age, gender, BMI, ethnicity, and geographic location of sample collection.

### Multivariable MR (MVMR)

MVMR is a conditional MR analysis technique^16^ that we used to determine whether the protective effect of serine was mediated via regulation of immune cell numbers; and whether the toxic effect of homostachydrine was detectable via coffee consumption or mediated via L-proline betaine. The p-value cut-offs used to choose instrumental SNPs for each exposure were chosen so as to achieve adequate instrument strength for both exposures (conditional F-statistic >10 for each exposure). Reported results showed no evidence of instrument heterogeneity (Cochran’s Q-test p > 0.05). Exposures were derived from independent cohorts and therefore a correction for the covariance between the effect of the genetic variants on each exposure was not necessary. MVMR was implemented using the MVMR (version 0.3) and MendelianRandomization (version 0.5.1) R packages.

### Rare variant analysis

MR is based on common genetic variants (MAF > 1%), which are largely orthogonal to large-effect missense rare variants. We therefore used the AstraZeneca PheWAS portal^17^ to analyse the ‘Union Primary Pulmonary Hypertension’ dataset (361,675 controls and 578 PAH cases) to determine whether rare variants within enzymes responsible for metabolism of our candidate metabolites are also associated with risk of PAH. Identified rare variants with a common biological effect were collapsed into a single Fisher’s exact two-sided test to determine whether the burden of variants is different in PAH cases and controls. We focused on LOF variants including protein-truncating variants, missense variants, and in-frame insertions and deletions which are damaging to the protein (REVEL score >=0.25)^18^ that were rare (MAF<0.001 if protein-truncating or MAF<0.0005 if non-protein-truncating, in both UK Biobank and GnomAD^19^); in addition we considered variants were high-quality variant-calls based on coverage, mapping quality, genotype quality, and Hardy-Weinberg equilibrium (‘ptvraredmg’ as described in^17^).

## Results

### Unbiased MR causally associates five metabolites with PAH risk

We hypothesised that serum metabolites may act upon pulmonary tissue to modify the biological processes leading to PAH. We used MR to perform a hypothesis-free study of the entire set of serum metabolites to determine which metabolites are causally related to PAH. To achieve this we obtained genetic variants associated with the serum concentration of 575 metabolites in 7,284 individuals^7,8^ (**Methods**). Next we applied a series of MR tests where the exposure was formed from metabolite-associated genetic variants, and genetic liability to PAH was the outcome measure (**Methods**). Genetic associations with PAH were determined within the FinnGen cohort consisting of 125 PAH patients and 162,837 controls^9^.

The complete results of our full unbiased metabolome screen are shown in **Supplementary Table 1**. After a stringent FDR correction of the IVW estimate p-value, five metabolites were significantly associated with risk of PAH. Two metabolites were protective against PAH: acetylphosphate (IVW p = 2.46e-06, beta = −16.06, se = 3.4) and serine (IVW p = 6.43e-06, beta = −6.7, se = 1.49, **Figure 2**, **Figure 3A-B**). Three metabolites increased the risk of PAH: homostachydrine (IVW p = 0.0003, beta = 1.09, se = 0.3), X-12029 (IVW p = 0.0006, beta = 8.9, se = 2.6) and X-11850 (IVW p = 0.0007, beta = 0.95, se = 0.28) (**Figure 2**, **Figure 3C-E**). None of these tests was invalidated by instrument pleiotropy or weak instruments (**Methods, Table 1**). Acetylphosphate was also significantly associated with PAH risk by the weighted median estimate (p = 0.04, beta =-13.7) and X-11850 was significant by the weighted mode estimate (p = 0.02, beta = 1.93). Only homostachydrine was significant in multiple robust MR estimates (weighted median p = 0.03, beta = 1.28; MR Egger p = 0.003, beta = 1.38; weighted mode p = 0.006, beta = 1.26, **Methods, Table 1**) which lends significant support to the validity of this metabolite as a mediator of toxicity leading to PAH.

**Figure 2:**
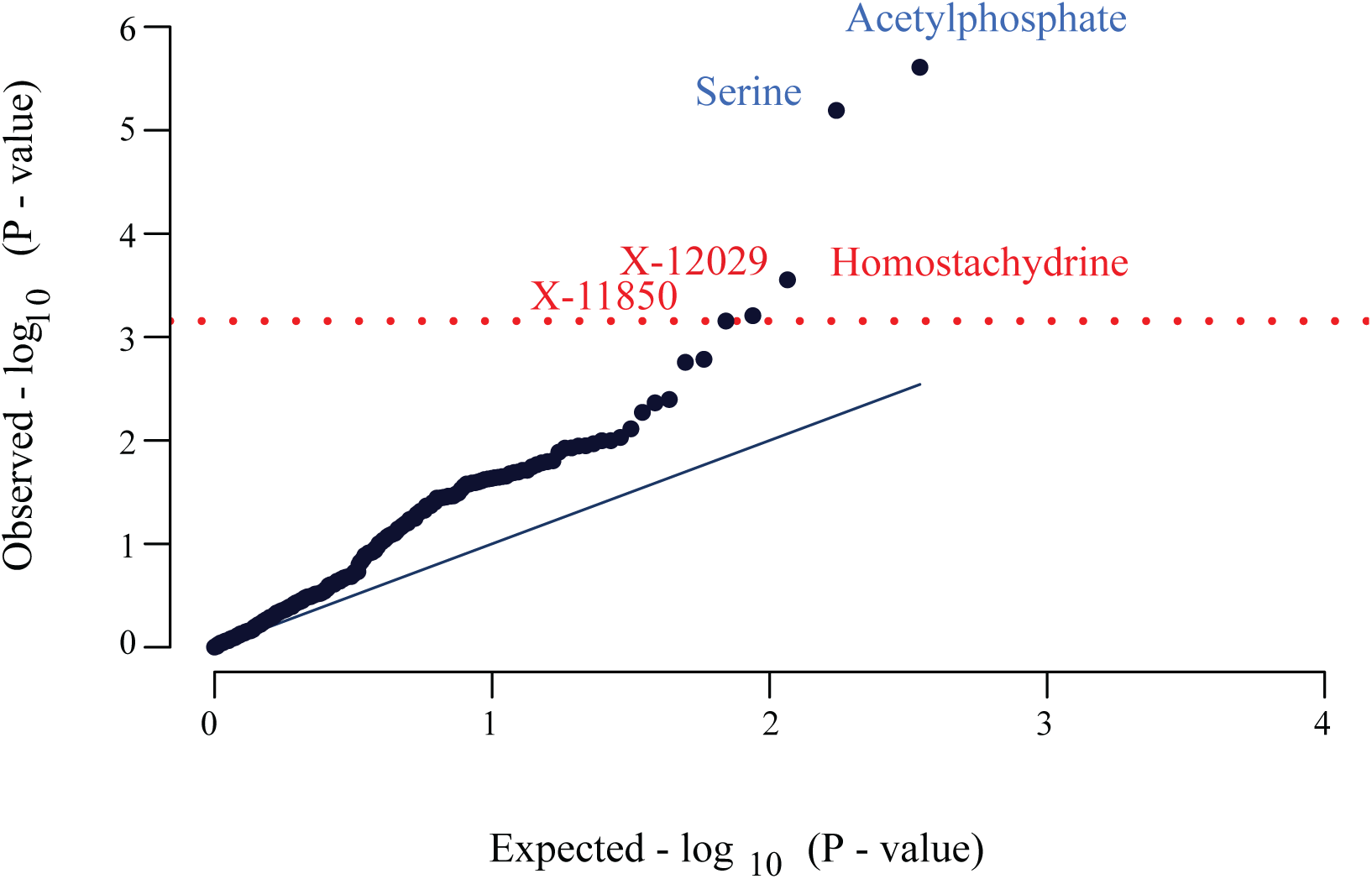
Five metabolites were significantly associated with risk of PAH by MR. QQ-plot of IVW p-values demonstrates that five metabolites passed a FDR multiple testing correction (red line). Blue text represents a protective association and red text represents a harmful association.

**Figure 3:**
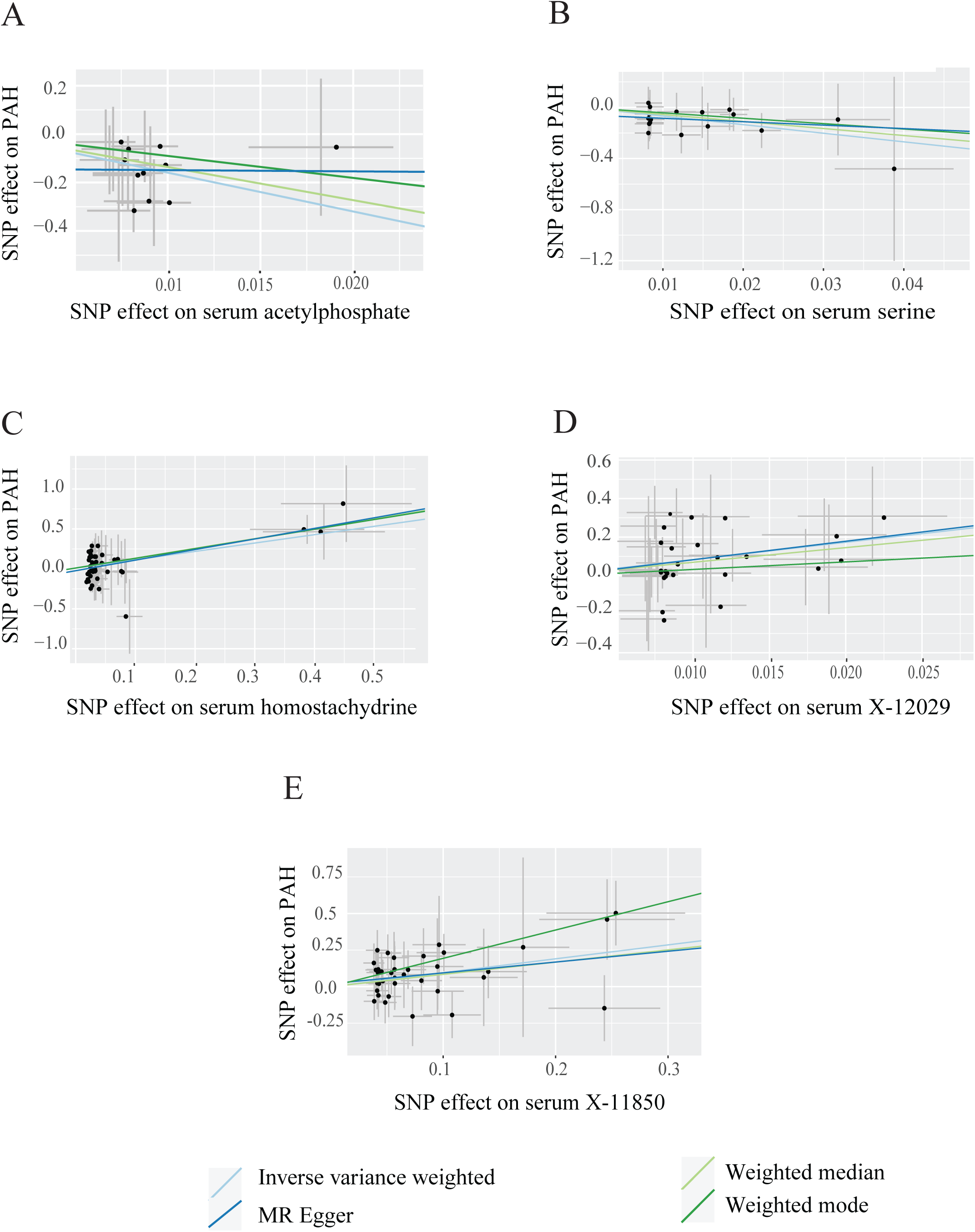
Two-sample MR tests for a causal effect of metabolites on risk of PAH. Scatter plots demonstrate a significant association of serum metabolite concentrations with risk of PAH including in robust MR tests for (A) acetylphosphate (B) serine, (C) homostachydrine (D) X-12029 and (E) X-11850. Each point represents the effect size (beta) and standard errors for each SNP-outcome relationship.

**Table 1:** Robust MR measures for the effect of significant metabolites on risk of PAH. Statistically significant results are highlighted in grey.

| Test | Serine | Acetylphosphate | Homostachydrine | X-12029 | X-11850 |
| --- | --- | --- | --- | --- | --- |
| IVW P Value | 6.43e-6 | 2.46e-06 | 0.0003 | 0.0006 | 0.0007 |
| IVW beta | -6.7 | -16.06 | 1.09 | 8.9 | 0.95 |
| Weighted Median P value | 0.13 | 0.04 | 0.03 | 0.07 | 0.1 |
| Weighted Median beta | -5.5 | -13.7 | 1.28 | 7.6 | 0.84 |
| Egger P value | 0.69 | 0.98 | 0.003 | 0.33 | 0.23 |
| Egger beta | -2.68 | -0.51 | 1.38 | 9.58 | 0.74 |
| Weighted Mode P value | 0.36 | 0.4 | 0.006 | 0.5 | 0.02 |
| Weighted Mode beta | -4.2 | -9.08 | 1.26 | 3.9 | 1.93 |
| Mean F test | 35 | 25 | 18 | 22 | 18 |
| IVW Cochran's Q test P value | 1 | 0.93 | 0.8 | 0.8 | 0.9 |
| Radial MR Outlier SNPs | 0 | 0 | 2 | 1 | 4 |
| Egger intercept test | 0.51 | 0.53 | 0.3 | 0.9 | 0.7 |
| I<sup>2</sup> | 0.97 | 0.96 | 0.94 | 0.96 | 0.95 |
| Number of SNPs LOO >0.05 | 0 | 0 | 0 | 0 | 0 |
| Total Number of SNPs | 17 | 11 | 42 | 25 | 36 |
| Number of Pleiotropic SNPs | 0 | 0 | 0 | 0 | 0 |

### Validation in a different PAH GWAS supports the validity of identified metabolites

To validate the set of metabolites we have associated with risk of PAH, we obtained a different larger FinnGen GWAS consisting of 208 PAH cases and 243,756 controls^9^ that has some overlap with our original sample set. Repeating the MR tests for the association of serum metabolite concentration with risk of PAH in this dataset confirmed our previous results for serine, acetylphosphate and homostachydrine but not for X-12029 and X-11850. In this analysis both serine (IVW p = 0.0004, beta = −5.23, se = 1.49) and acetylphosphate (IVW p = 0.0026, beta = −11.3, se = 3.76) were significantly protective against PAH while homostachydrine increased risk of PAH (IVW p = 0.004, beta = 0.67, se = 0.24, **Supplementary Figure S1)**. X-12029 (IVW P = 0.1, beta = 3.6 se = 2.2) and X-11850 (IVW P = 0.056, beta = 0.5 se = 0.26) were not significantly associated with the increased risk of PAH but maintained the same direction of effect. After this result X-12029 and X-11850 were excluded from further analysis as these results were deemed to be non-reproducible.

### Direct measurement of serine and homostachydrine supports a link to PAH clinical severity

MR is a powerful tool that leverages large sample sets. However, MR relies on genetic correlates of measured exposures and outcomes which is not equivalent to a direct measurement. We therefore sought to further validate our results by direct measurement of metabolites in a set of clinically profiled PAH patients from the PAH UK data of 449 PAH patients including 260 idiopathic PAH patients^5,6^. Unlike our MR analysis, this experiment cannot exclude reverse causality, however where direct measurement supports our MR analyses we conclude that there is convergent evidence that a particular metabolite modifies biological processes leading to PAH.

In this clinical dataset, the plasma concentration of both serine and homostachydrine were significantly correlated with multiple measures of PAH severity after adjusting for age, gender, BMI and geographical location (**Methods**, **Table 2**). In each case the direction of effect was consistent with our MR analyses i.e. serine was protective and homostachydrine was harmful. In addition, serum homostachydrine was significantly correlated with a reduction in 5 year survival (Cox regression, cor= 6.76e-02, p= 0.03, **Methods**) after adjusting for age, gender, site and ethnicity **(Figure 4)**. Serum serine concentration was correlated with an increase in survival but after adjusting for covariates, the results were not significant **(Supplementary Figure S2)**. Direct measurement of acetylphosphate was not possible and therefore there was no clinical evidence to support or refute the role of acetylphosphate in PAH; to be cautious we excluded acetylphosphate from further analysis.

**Figure 4:**
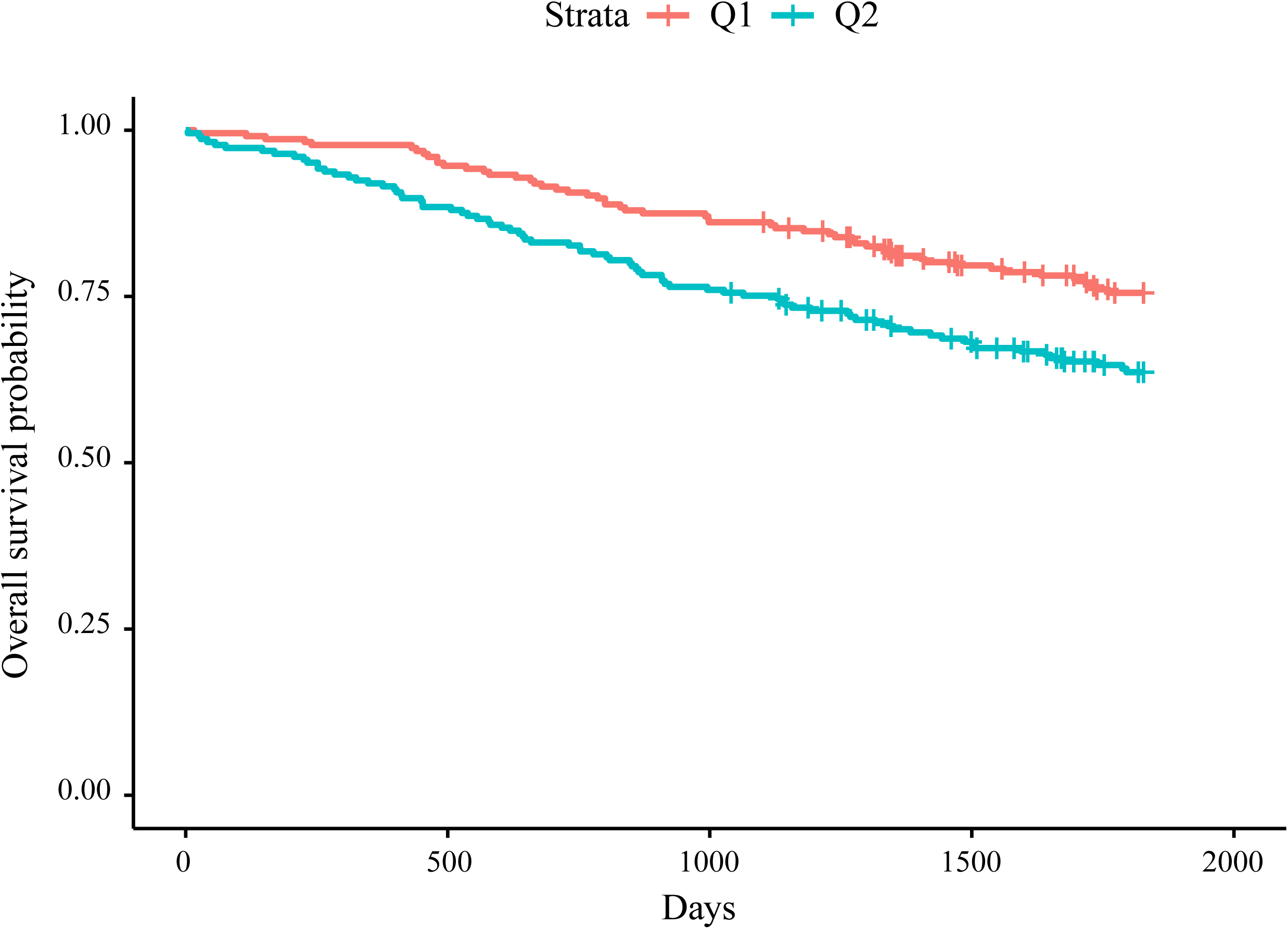
Higher serum homostachydrine is associated with a significant reduction in five year survival rate in the UK PAH cohort. (Cox regression, cor= 6.76e-02, p= 0.03) after adjusting for age, gender, BMI, geographical site and ethnicity. Q1: < 50% quantile, Q2: > 50% quantile.

**Table 2:** Linear and Logistic regression of biomarkers of PAH severity against plasma concentration of metabolites. All tests were adjusted for age, gender, BMI and geographical location. Statistically significant results are highlighted in grey.

| Variable | Serine |  | Homostachydrine |  |
| --- | --- | --- | --- | --- |
|  | coefficient | p-value | coefficient | p-value |
| **WHO-FC** | -3.9 | 0.72 | 1.66 | 0.56 |
| **NT-proBNP** | -0.5 | 0.22 | 0.035 | 0.05 |
| **6MWD** | -0.5 | 0.22 | -2.15 | 0.07 |
| **mPAP** | -2 | 0.45 | 0.19 | 0.09 |
| **PAWP** | -0.05 | 0.94 | 0.069 | 0.047 |
| **SvO2** | 2.5 | 0.27 | -0.21 | 0.025 |
| **FVCp** | 12.4 | 0.009 | -0.3 | 0.11 |
| **Cardiac Index** | 0.56 | 0.005 | -0.01 | 0.19 |
| **Cardiac Output** | 0.73 | 0.027 | -0.017 | 0.2 |
| **PVRdynes** | -72 | 0.48 | 8.27 | 0.049 |
| **REVEAL Scores** | -4.9 | 0.71 | 0.09 | 0.72 |

The WHO functional class (WHO-FC), N-terminal pro-brain natriuretic peptide (NT- proBNP), Six-minute walking distance (6MWD), mean pulmonary arterial pressure (mPAP), pulmonary arterial wedge pressure (PAWP), Venous oxygen saturation (SvO2), percentage of predicted forced vital capacity (FVCp), Cardiac index (CI) , cardiac output (CO) and pulmonary vascular resistance in dynes (PVRdynes).

### Homostachydrine association with PAH is independent of coffee consumption

Homostachydrine is a xenobiotic metabolite (also known as pipecolic acid betaine) that is highly homologous to L-proline betaine (also called stachydrine). Initially, it was discovered in leaves of Medicago (Alfalfa) and Achillea genera. Homostachydrine is also present within frequently utilised Robusta coffee beans^20^. Indeed, serum homostachydrine has been proposed as a marker of coffee consumption^21^. We have demonstrated that serum homostachydrine is causally associated with PAH risk and severity, however it may not be directly related to PAH. Instead it may act via an intermediate which is also present within coffee. To investigate this possibility we performed a MR test for the effect of coffee consumption on risk of PAH.

We utilised a GWAS for questionnaire-reported coffee consumption consisting of 428,860 individuals^22^. There was no evidence of a significant causal effect of coffee consumption on risk of PAH (IVW p=0.87, beta= 0.27). In addition, we performed a conditional analysis of the effect of homostachydrine on risk of PAH independent of coffee consumption using MVMR (**Methods**), MVMR revealed that the association of homostachydrine with PAH was not invalidated when conditioned on coffee consumption (beta = 0.97, p=0.006). These data suggest that homostachydrine is acting directly and is not serving as a proxy for another component of coffee.

### The effect of L-proline betaine effect on PAH is dependant on homostachydrine

Homostachydrine is structurally similar to L-proline betaine but it contains a different alkyl chain length and substitution pattern. It is likely that the two share a common metabolic pathway and their serum concentrations may be correlated. Interestingly, an increase in serum L-proline betaine has been associated with a poor prognosis in PAH^23^. Our MR analysis supports this conclusion: We found a borderline significant toxic effect of L-proline betaine on PAH (IVW p = 0.055, beta =5.14, se =2.68). This raised a question as to whether the effects of homostachydrine and L-proline betaine are independent. To investigate this further we performed a MVMR (**Methods**) in which we discovered that the effect of L-proline betaine is removed (beta = 1.8, p=0.39) when we conditioned on homostachydrine; whereas homostachydrine remained significantly associated with increased risk of PAH (beta = 2.04, p=0.054) even when conditioned on L-proline betaine. We conclude that the effect of homostachydrine on risk of PAH is independent of the effect of L-proline betaine; and homostachydrine may even account for previously reported observations regarding the effect of L-proline betaine on PAH.

### There is no MR evidence that the effect of serine on PAH is mediated via immunomodulation

We have identified a protective effect of serine on risk and severity of PAH. We interrogated the literature to determine a likely mechanism for this effect. Activated immune cells are rapidly proliferating and are thus dependent on serine to function^24^. Moreover, PAH patients exhibit immune system abnormalities^25–27^. We hypothesised that the protective effect of serine on PAH may be mediated via immunomodulation.

To investigate this further we performed a second unbiased MR screen of immune traits ^11^ as determinants of PAH risk (**Supplementary Table 2, Methods**). After FDR multiple testing correction, only two traits were significantly associated with risk of PAH: the proportion of Vδ1+ T cells (IVW, p=0.00006, beta=-1.02) and the proportion of CD1c-myeloid dendritic cells (IVW, p=0.00076, beta= 0.84) but serine was not significantly associated with either trait (IVW, p>0.05). Nineteen immune traits were nominally (IVW p<0.05) associated with PAH risk (**Supplementary Table 3, Methods**); but only two of these immune traits: the concentration of CD8+ T cells (IVW, p=0.013, beta=0.6), and the concentration of CD3 on double negative (DN) T cells (IVW, p=0.042, beta=0.9), were also significantly associated with serum concentration of serine. We applied MVMR to determine whether the effect of serine on PAH was mediated via either of these immune traits; but on the contrary, in MVMR (**Methods**) the effect of serine on PAH remained significant (p<0.05) but both immune traits were non-significant when conditioned on serine. Our MR analysis thereby suggests that the effect of serine is independent of immune function and not mediated via CD8 T cells or CD3+ DN T-cells.

### Rare variant analysis supports the validity of serine as protective against PAH

If serum serine is protective against PAH it follows that genetic variants which reduce synthesis of endogenous serine might modify the risk of PAH. ATF4 is a transcription factor which promotes the expression of enzymes involved in endogenous synthesis of serine under conditions of serine starvation^28^ and has previously been linked to a related disease, dilated cardiomyopathy (DCM)^29^. We performed rare variant genetic burden testing (**Methods**) in a cohort of 578 PAH cases compared to n = *361,675* controls to determine whether PAH patients carried an excess of LOF mutations within ATF4 or any of the enzymes under its control (**Table 3**). There was an excess of loss-of-function (LOF) mutations within ATF4 in PAH cases compared to controls (Fisher’s exact test, OR=3.78, p=0.047), which is consistent with our MR data suggesting a protective role for serum serine. There was no evidence for an excess of LOF variants in any of the individual enzymes involved in serine biosynthesis

**Table 3:** Loss of function (LOF) mutations within the ATF4 transcription factor involved in control of serine biosynthesis are associated with increased risk of PAH. This analysis utilised whole exome sequencing from 578 PAH patients and 361,675 controls ^17^. Numbers refer to the total number of PAH cases and controls who carried an LOF variant under the ‘ptvraredmg’ classification^17^. Statistically significant results are highlighted in grey.

| Gene | P value | Number of cases | Number of controls | OR | Biological role |
| --- | --- | --- | --- | --- | --- |
| **ATF4** | 0.047 | 3 (0.53%) | 498 (0.14%) | 3.78 | TF which upregulates expression of PSPH, PSAT, PHGDH and SHMT2 |
| **PSPH** | >0.1 | - | - | - | Serine biosynthesis |
| **PSAT** | >0.1 | - | - | - |  |
| **PHGDH** | >0.1 | - | - | - |  |
| **SHMT2** | >0.1 | - | - | - | Conversion between glycine and serine |

## Discussion

PAH is an archetypal complex disease determined by gene-environment interactions which is currently incurable. The role of the metabolome in the development of PAH is poorly understood although groups of metabolites have been observed to associate with PAH risk and severity in previous observational studies^5^. Here we have applied an unbiased approach to search for causal links between serum metabolites and PAH risk, leveraging the significant statistical power provided by two-sample MR. We have previously used MR to perform an unbiased search for metabolites which determine risk for amyotrophic lateral sclerosis (ALS)^12^, Alzheimer’s disease (AD)^30^ and age-related macular degeneration^31^.

We have linked three metabolites to the risk of PAH using MR in two different GWAS for PAH: acetylphosphate, serine and homostachydrine. Serine and homostachydrine are supported by orthogonal analysis via direct measurement of serum metabolites in a clinical cohort of PAH patients. In our MR results, higher serum serine reduces risk for PAH and in the clinical cohort higher serum serine was associated with reduced PAH severity and increased survival. In contrast, our MR results higher serum homostachydrine increased risk for PAH, and in the clinical cohort higher serum homostachydrine was associated with increased PAH severity and reduced survival. The orthogonal nature of the tests we have applied provides significant evidence that we have identified new biological mechanisms underpinning PAH.

Serine biosynthesis has been previously implicated in dilated cardiomyopathy (DCM) via the transcription factor ATF4^29^; DCM and PAH may share underlying pathobiology, particularly related to vascular remodelling^32^. Our rare variant analysis associated LOF mutations within ATF4 with increased risk of PAH which supports a role for this pathway in PAH. In the study of the effect of serine biosynthesis on DCM^29^, positive treatment effects were correlated with correction of mitochondrial respiration chain defects suggesting a boost to energy production. Our analyses suggest that serine is not linked directly to PAH via immunomodulation and we conclude that a role in energy metabolism is more likely. Promoting endogenous serine synthesis via the activation of ATF4 may be a novel therapeutic strategy for PAH.

Homostachydrine is a xenobiotic metabolite which is exclusively produced in plants including within Robusta coffee beans. Homostachydrine has been proposed as a marker of quality for coffee which is thought to be improved by a higher proportion of Arabica coffee beans^20^. Indeed, serum homostachydrine has been proposed as a marker of coffee consumption^21^. However our analysis suggests that the effect of homostachydrine on PAH is independent of coffee consumption and therefore not likely to be a byproduct of some other component of coffee.

Homostachydrine is structurally similar to L-proline betaine and our MR analysis suggests that L-proline betaine may be indirectly linked to risk of PAH via homostachydrine. Higher fasting blood concentrations of L-proline betaine have been linked to poor prognosis in PAH, including a rise in NT-proBNP which is an important biomarker of PAH severity^23^. Unlike homostachydrine there is some understanding of the specific activities and physiological roles of L-proline betaine, for example increased concentrations of L-proline betaine may disrupt mitochondrial structure and function leading to a toxic energy deficit^33^. It is possible that this effect on energy metabolism is actually mediated via homostachydrine, and that an energy deficit is responsible for the link between serum homostachydrine and the risk and severity of PAH.

In conclusion we have demonstrated the utility of hypothesis-free MR for the discovery of disease-associated metabolites. Our follow-up analysis in a clinical cohort adds significant orthogonal evidence for our conclusions by adding a direct measurement of serum metabolites in PAH patients. We conclude that there is consistent evidence for a protective effect of serum serine on PAH and a harmful effect of homostachydrine. Both of these metabolites may be acting via modulation of energy metabolism. Further study is necessary and could lead to guidance cautioning consumption of certain food-products containing homostachydrine, or increasing serine consumption, for PAH patients.

## Author contributions

**Elham Alhathli:** Conceptualization, Data curation, Formal analysis, Investigation, Methodology, Validation, Visualization and Writing - original draft. **Johnathan Cooper-Knock:** Conceptualization, Project administration, Resources, Software, Supervision, Funding acquisition, review & editing. **Dennis Wang:** Supervision, review & editing. **Stefan Gräf:** Project administration, Resources, review & editing. **Niamh Errington**: Data curation, Formal analysis, Investigation, Methodology, review & editing. **A A Roger Thompson:** Supervision, review & editing. **Allan Lawrie:** Supervision, review & editing **Thomas H Julian:** Conceptualization, Data curation, Formal analysis and Methodology. **Zain Ul Abideen Girach:** Data Visualization and Review. **Christopher Rhodes:** Project administration, Resources, Formal analysis, Supervision, review & editing. **Martin Wilkins:** Project administration, Resources, Supervision, review & editing.

## Disclosure statement

All authors report no conflict of interest.

## Data Availability

All data used in this manuscript is publicly available via citations in the manuscript

## Acknowledgements

This work was supported by a Wellcome Trust fellowship [216596/Z/19/Z to J.C.-K]. AL is supported by a British Heart Foundation Senior Basic Science Fellowship [FS/18/52/33808]. CJR is supported by BHF Basic Science Research fellowships (FS/15/59/31839 & FS/SBSRF/21/31025) and Academy of Medical Sciences Springboard fellowship (SBF004\1095). THJ is supported by the NIHR academic clinical fellowship. AART was supported by a BHF Intermediate Clinical Fellowship (FS/18/13/33281). This work was supported by the NIHR BioResource which supports the UK National Cohort of Idiopathic and Heritable PAH; the British Heart Foundation (BHF SP/12/12/29836) and the UK Medical Research Council (MR/K020919/1). This work was supported in part by the British Heart Foundation Centre for Research Excellence award RE/18/4/34215. We acknowledge the UK National Cohort Study of Idiopathic and Heritable PAH. We thank National Institute for Health Research (NIHR) BioResource volunteers for their participation, and gratefully acknowledge NIHR BioResource centres, NHS Trusts and staff for their contribution. We thank the NIHR Imperial Clinical Research Facility, NIHR Sheffield Biomedical Research Centre, Cambridge NIHR Cardiorespiratory BRC and NHS Blood and Transplant. The views expressed are those of the author(s) and not necessarily those of the NHS, the NIHR or the Department of Health and Social Care.

---

**Supplementary Table 1:** Unbiased two-sample MR metabolites screen results for 560 metabolites as exposures for PAH risk with all the sensitivity measures and robust tests results.

**Supplementary Table 2:** Unbiased two-sample MR immune traits screens for 144 traits as exposures for PAH risk with all the sensitivity measures and robust tests results.

**Supplementary Table 3:** IVW two sample MR screen with serine as exposure and significant immune traits against the risk of PAH as output (19 immune traits)

**Supplementary Figure 1:**
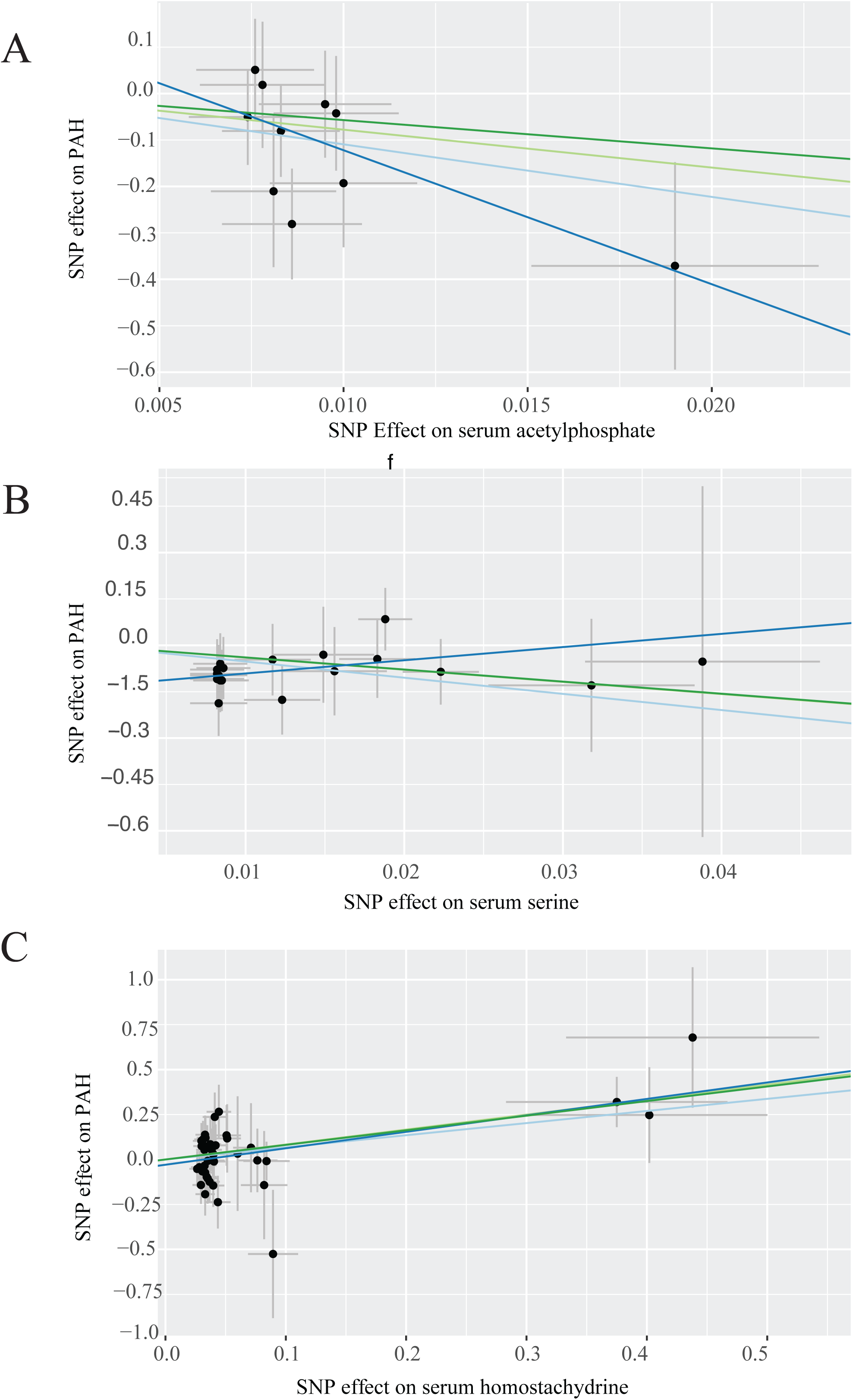
Two-sample MR tests for a causal effect of metabolites on risk of PAH in a validation GWAS. Scatter plots demonstrate a significant association of serum metabolite concentrations with risk of PAH including in robust MR tests for (A) acetylphosphate (B) serine, and (C) homostachydrine. Each point represents the effect size (beta) and standard errors for each SNP-outcome relationship

**Supplementary Figure 2:**
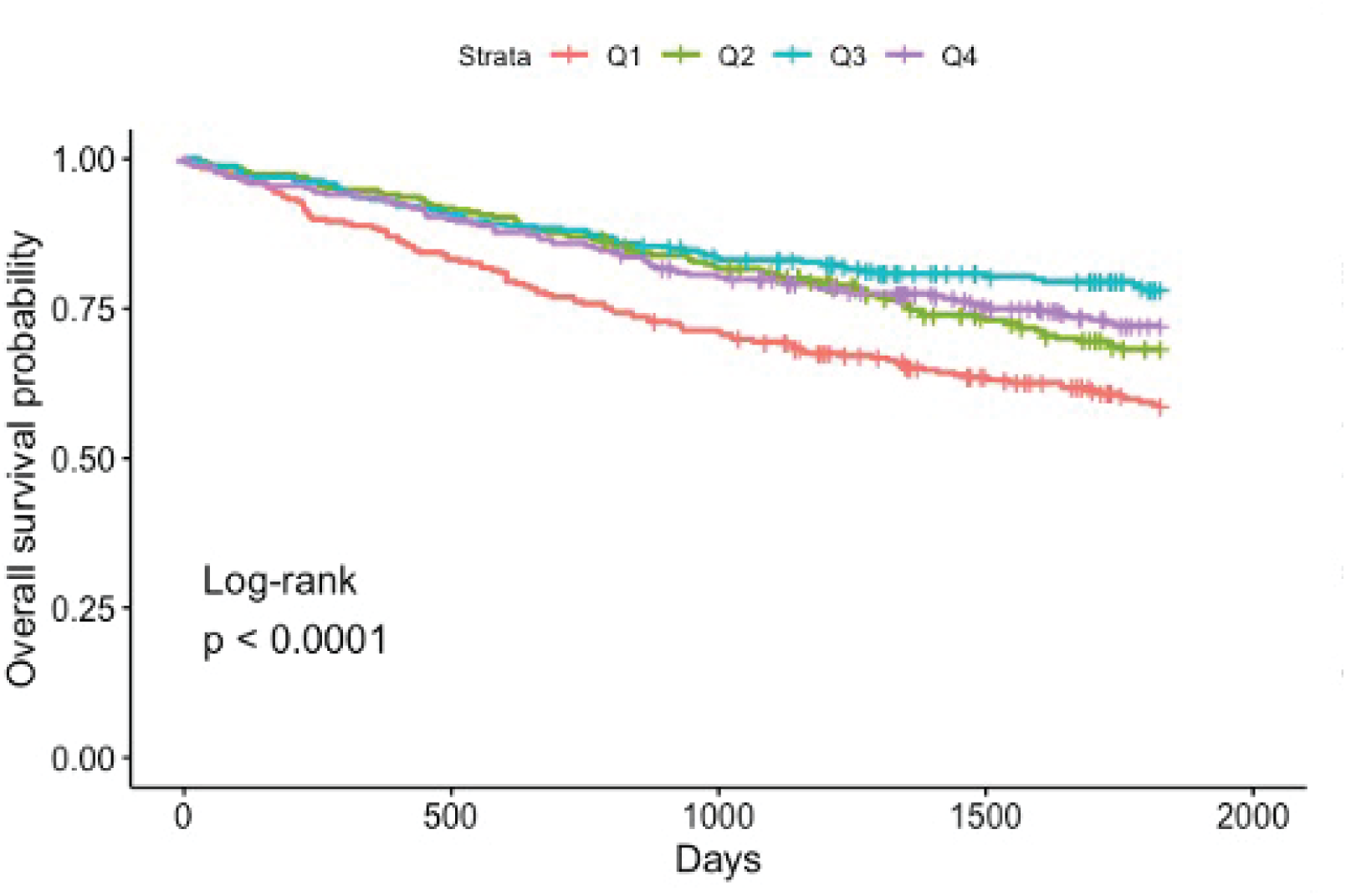
Lower serum serine is associated with a significant reduction in five year survival rate in the UK PAH cohort. (Cox regression, cor= 6.76e-02, p= 0.03) after adjusting for age, gender, bmi, site and ethnicity. Q1: < 25% quantile, Q2: < 50% quantile; Q3: < 75% quantile, < 100% quantile

## Notes

### Competing Interest Statement

The authors have declared no competing interest.

## References

1. Rhodes CJ, Sweatt AJ, Maron BA. Harnessing Big Data to Advance Treatment and Understanding of Pulmonary Hypertension. Circ Res. 2022;130:1423–1444.

2. Lemay S-E, Awada C, Shimauchi T, Wu W-H, Bonnet S, Provencher S, Boucherat O. Fetal Gene Reactivation in Pulmonary Arterial Hypertension: GOOD, BAD, or BOTH? Cells [Internet]. 2021;10. Available from: http://dx.doi.org/10.3390/cells10061473

3. Julian TH, Boddy S, Islam M, Kurz J, Whittaker KJ, Moll T, Harvey C, Zhang S, Snyder MP, McDermott C, Cooper-Knock J, Shaw PJ. A review of Mendelian randomization in amyotrophic lateral sclerosis. Brain. 2022;145:832–842.

4. Davey Smith G, Ebrahim S. “Mendelian randomization”: can genetic epidemiology contribute to understanding environmental determinants of disease? Int J Epidemiol. 2003;32:1–22.

5. Rhodes CJ, Ghataorhe P, Wharton J, Rue-Albrecht KC, Hadinnapola C, Watson G, Bleda M, Haimel M, Coghlan G, Corris PA, Howard LS, Kiely DG, Peacock AJ, Pepke-Zaba J, Toshner MR, Wort SJ, Gibbs JSR, Lawrie A, Gräf S, Morrell NW, Wilkins MR. Plasma Metabolomics Implicates Modified Transfer RNAs and Altered Bioenergetics in the Outcomes of Pulmonary Arterial Hypertension. Circulation. 2017;135:460–475.

6. Swietlik EM, Ghataorhe P, Zalewska KI, Wharton J, Howard LS, Taboada D, Cannon JE, UK National Cohort Study of PAH, Morrell NW, Wilkins MR, Toshner M, Pepke-Zaba J, Rhodes CJ. Plasma metabolomics exhibit response to therapy in chronic thromboembolic pulmonary hypertension. Eur Respir J [Internet]. 2021;57. Available from: http://dx.doi.org/10.1183/13993003.03201-2020

7. Shin S-Y, Fauman EB, Petersen A-K, Krumsiek J, Santos R, Huang J, Arnold M, Erte I, Forgetta V, Yang T-P, Walter K, Menni C, Chen L, Vasquez L, Valdes AM, Hyde CL, Wang V, Ziemek D, Roberts P, Xi L, Grundberg E, Multiple Tissue Human Expression Resource (MuTHER) Consortium, Waldenberger M, Richards JB, Mohney RP, Milburn MV, John SL, Trimmer J, Theis FJ, Overington JP, Suhre K, Brosnan MJ, Gieger C, Kastenmüller G, Spector TD, Soranzo N. An atlas of genetic influences on human blood metabolites. Nat Genet. 2014;46:543–550.

8. Kettunen J, Demirkan A, Würtz P, Draisma HHM, Haller T, Rawal R, Vaarhorst A, Kangas AJ, Lyytikäinen L-P, Pirinen M, Pool R, Sarin A-P, Soininen P, Tukiainen T, Wang Q, Tiainen M, Tynkkynen T, Amin N, Zeller T, Beekman M, Deelen J, van Dijk KW, Esko T, Hottenga J-J, van Leeuwen EM, Lehtimäki T, Mihailov E, Rose RJ, de Craen AJM, Gieger C, Kähönen M, Perola M, Blankenberg S, Savolainen MJ, Verhoeven A, Viikari J, Willemsen G, Boomsma DI, van Duijn CM, Eriksson J, Jula A, Järvelin M-R, Kaprio J, Metspalu A, Raitakari O, Salomaa V, Slagboom PE, Waldenberger M, Ripatti S, Ala-Korpela M. Genome-wide study for circulating metabolites identifies 62 loci and reveals novel systemic effects of LPA. Nat Commun. 2016;7:11122.

9. Kurki MI, Karjalainen J, Palta P, Sipilä TP, Kristiansson K, Donner KM, Reeve MP, Laivuori H, Aavikko M, Kaunisto MA, Loukola A, Lahtela E, Mattsson H, Laiho P, Della Briotta Parolo P, Lehisto AA, Kanai M, Mars N, Rämö J, Kiiskinen T, Heyne HO, Veerapen K, Rüeger S, Lemmelä S, Zhou W, Ruotsalainen S, Pärn K, Hiekkalinna T, Koskelainen S, Paajanen T, Llorens V, Gracia-Tabuenca J, Siirtola H, Reis K, Elnahas AG, Sun B, Foley CN, Aalto-Setälä K, Alasoo K, Arvas M, Auro K, Biswas S, Bizaki-Vallaskangas A, Carpen O, Chen C-Y, Dada OA, Ding Z, Ehm MG, Eklund K, Färkkilä M, Finucane H, Ganna A, Ghazal A, Graham RR, Green EM, Hakanen A, Hautalahti M, Hedman ÅK, Hiltunen M, Hinttala R, Hovatta I, Hu X, Huertas-Vazquez A, Huilaja L, Hunkapiller J, Jacob H, Jensen J-N, Joensuu H, John S, Julkunen V, Jung M, Junttila J, Kaarniranta K, Kähönen M, Kajanne R, Kallio L, Kälviäinen R, Kaprio J, FinnGen, Kerimov N, Kettunen J, Kilpeläinen E, Kilpi T, Klinger K, Kosma V-M, Kuopio T, Kurra V, Laisk T, Laukkanen J, Lawless N, Liu A, Longerich S, Mägi R, Mäkelä J, Mäkitie A, Malarstig A, Mannermaa A, Maranville J, et al. FinnGen provides genetic insights from a well-phenotyped isolated population. Nature. 2023;613:508–518.

10. Marshall DD, Powers R. Beyond the paradigm: Combining mass spectrometry and nuclear magnetic resonance for metabolomics. Prog Nucl Magn Reson Spectrosc. 2017;100:1–16.

11. Roederer M, Quaye L, Mangino M, Beddall MH, Mahnke Y, Chattopadhyay P, Tosi I, Napolitano L, Terranova Barberio M, Menni C, Villanova F, Di Meglio P, Spector TD, Nestle FO. The genetic architecture of the human immune system: a bioresource for autoimmunity and disease pathogenesis. Cell. 2015;161:387–403.

12. Boddy S, Islam M, Moll T, Kurz J, Burrows D, McGown A, Bhargava A, Julian TH, Harvey C, Marshall JN, Hall BP, Allen SP, Kenna KP, Sanderson E, Zhang S, Ramesh T, Snyder MP, Shaw PJ, McDermott C, Cooper-Knock J. Unbiased metabolome screen leads to personalized medicine strategy for amyotrophic lateral sclerosis. Brain Commun. 2022;4:fcac069.

13. Burgess S, Butterworth A, Thompson SG. Mendelian randomization analysis with multiple genetic variants using summarized data. Genet Epidemiol. 2013;37:658–665.

14. Bowden J, Spiller W, Del Greco M F, Sheehan N, Thompson J, Minelli C, Davey Smith G. Improving the visualization, interpretation and analysis of two-sample summary data Mendelian randomization via the Radial plot and Radial regression. Int J Epidemiol. 2018;47:1264–1278.

15. Benza RL, Gomberg-Maitland M, Elliott CG, Farber HW, Foreman AJ, Frost AE, McGoon MD, Pasta DJ, Selej M, Burger CD, Frantz RP. Predicting Survival in Patients With Pulmonary Arterial Hypertension: The REVEAL Risk Score Calculator 2.0 and Comparison With ESC/ERS-Based Risk Assessment Strategies. Chest. 2019;156:323–337.

16. Burgess S, Thompson SG. Multivariable Mendelian randomization: the use of pleiotropic genetic variants to estimate causal effects. Am J Epidemiol. 2015;181:251–260.

17. Wang Q, Dhindsa RS, Carss K, Harper AR, Nag A, Tachmazidou I, Vitsios D, Deevi SVV, Mackay A, Muthas D, Hühn M, Monkley S, Olsson H, AstraZeneca Genomics Initiative, Wasilewski S, Smith KR, March R, Platt A, Haefliger C, Petrovski S. Rare variant contribution to human disease in 281,104 UK Biobank exomes. Nature. 2021;597:527–532.

18. Ioannidis NM, Rothstein JH, Pejaver V, Middha S, McDonnell SK, Baheti S, Musolf A, Li Q, Holzinger E, Karyadi D, Cannon-Albright LA, Teerlink CC, Stanford JL, Isaacs WB, Xu J, Cooney KA, Lange EM, Schleutker J, Carpten JD, Powell IJ, Cussenot O, Cancel-Tassin G, Giles GG, MacInnis RJ, Maier C, Hsieh C-L, Wiklund F, Catalona WJ, Foulkes WD, Mandal D, Eeles RA, Kote-Jarai Z, Bustamante CD, Schaid DJ, Hastie T, Ostrander EA, Bailey-Wilson JE, Radivojac P, Thibodeau SN, Whittemore AS, Sieh W. REVEL: An Ensemble Method for Predicting the Pathogenicity of Rare Missense Variants. Am J Hum Genet. 2016;99:877–885.

19. Chen S, Francioli LC, Goodrich JK, Collins RL, Kanai M, Wang Q, Alföldi J, Watts NA, Vittal C, Gauthier LD, Poterba T, Wilson MW, Tarasova Y, Phu W, Yohannes MT, Koenig Z, Farjoun Y, Banks E, Donnelly S, Gabriel S, Gupta N, Ferriera S, Tolonen C, Novod S, Bergelson L, Roazen D, Ruano-Rubio V, Covarrubias M, Llanwarne C, Petrillo N, Wade G, Jeandet T, Munshi R, Tibbetts K, gnomAD Project Consortium, O’Donnell-Luria A, Solomonson M, Seed C, Martin AR, Talkowski ME, Rehm HL, Daly MJ, Tiao G, Neale BM, MacArthur DG, Karczewski KJ. A genome-wide mutational constraint map quantified from variation in 76,156 human genomes [Internet]. bioRxiv. 2022 [cited 2023 Jun 7];2022.03.20.485034. Available from: https://www.biorxiv.org/content/biorxiv/early/2022/10/10/2022.03.20.485034

20. Servillo L, Giovane A, Casale R, Cautela D, D’Onofrio N, Balestrieri ML, Castaldo D. Homostachydrine (pipecolic acid betaine) as authentication marker of roasted blends of Coffea arabica and Coffea canephora (Robusta) beans. Food Chem. 2016;205:52–57.

21. He WJ, Chen J, Razavi AC, Hu EA, Grams ME, Yu B, Parikh CR, Boerwinkle E, Bazzano L, Qi L, Kelly TN, Coresh J, Rebholz CM. Metabolites Associated with Coffee Consumption and Incident Chronic Kidney Disease. Clin J Am Soc Nephrol. 2021;16:1620–1629.

22. Elsworth B, Lyon M, Alexander T, Liu Y, Matthews P, Hallett J, Bates P, Palmer T, Haberland V, Smith GD, Zheng J, Haycock P, Gaunt TR, Hemani G. The MRC IEU OpenGWAS data infrastructure [Internet]. bioRxiv. 2020 [cited 2023 Jun 8];2020.08.10.244293. Available from: https://www.biorxiv.org/content/10.1101/2020.08.10.244293

23. Yang Y, Xu J, Zhou J, Xue J, Gao J, Li X, Sun B, Yang B, Liu Z, Zhao Z, Luo Q, Zeng Q, Zheng L, Xiong C. High Betaine and Dynamic Increase of Betaine Levels Are Both Associated With Poor Prognosis of Patients With Pulmonary Hypertension. Front Cardiovasc Med. 2022;9:852009.

24. Wu Q, Chen X, Li J, Sun S. Serine and Metabolism Regulation: A Novel Mechanism in Antitumor Immunity and Senescence. Aging Dis. 2020;11:1640–1653.

25. Tomaszewski M, Bębnowska D, Hrynkiewicz R, Dworzyński J, Niedźwiedzka-Rystwej P, Kopeć G, Grywalska E. Role of the Immune System Elements in Pulmonary Arterial Hypertension. J Clin Med Res [Internet]. 2021;10. Available from: http://dx.doi.org/10.3390/jcm10163757

26. Ni S, Ji T, Dong J, Chen F, Feng H, Zhao H, Chen D, Ma W. Immune Cells in Pulmonary Arterial Hypertension. Heart Lung Circ. 2022;31:934–943.

27. Jones RJ, De Bie EMDD, Groves E, Zalewska KI, Swietlik EM, Treacy CM, Martin JM, Polwarth G, Li W, Guo J, Baxendale HE, Coleman S, Savinykh N, Coghlan JG, Corris PA, Howard LS, Johnson MK, Church C, Kiely DG, Lawrie A, Lordan JL, Mackenzie Ross RV, Pepke Zaba J, Wilkins MR, Wort SJ, Fiorillo E, Orrù V, Cucca F, Rhodes CJ, Gräf S, Morrell NW, McKinney EF, Wallace C, Toshner M, UK National Cohort Study of Idiopathic and Heritable PAH Consortium. Autoimmunity Is a Significant Feature of Idiopathic Pulmonary Arterial Hypertension. Am J Respir Crit Care Med. 2022;206:81–93.

28. Tajan M, Hennequart M, Cheung EC, Zani F, Hock AK, Legrave N, Maddocks ODK, Ridgway RA, Athineos D, Suárez-Bonnet A, Ludwig RL, Novellasdemunt L, Angelis N, Li VSW, Vlachogiannis G, Valeri N, Mainolfi N, Suri V, Friedman A, Manfredi M, Blyth K, Sansom OJ, Vousden KH. Serine synthesis pathway inhibition cooperates with dietary serine and glycine limitation for cancer therapy. Nat Commun. 2021;12:366.

29. Perea-Gil I, Seeger T, Bruyneel AAN, Termglinchan V, Monte E, Lim EW, Vadgama N, Furihata T, Gavidia AA, Arthur Ataam J, Bharucha N, Martinez-Amador N, Ameen M, Nair P, Serrano R, Kaur B, Feyen DAM, Diecke S, Snyder MP, Metallo CM, Mercola M, Karakikes I. Serine biosynthesis as a novel therapeutic target for dilated cardiomyopathy. Eur Heart J. 2022;43:3477–3489.

30. Şanlı BA, Whittaker KJ, Motsi GK, Shen E, Julian TH, Cooper-Knock J. Unbiased metabolome screen links serum urate to risk of Alzheimer’s disease. Neurobiol Aging. 2022;120:167–176.

31. Julian TH, Cooper-Knock J, MacGregor S, Guo H, Aslam T, Sanderson E, Black GCM, Sergouniotis PI. Phenome-wide Mendelian randomisation analysis identifies causal factors for age-related macular degeneration. Elife [Internet]. 2023;12. Available from: http://dx.doi.org/10.7554/eLife.82546

32. Zhao Y-Y, Liu Y, Stan R-V, Fan L, Gu Y, Dalton N, Chu P-H, Peterson K, Ross J Jr, Chien KR. Defects in caveolin-1 cause dilated cardiomyopathy and pulmonary hypertension in knockout mice. Proc Natl Acad Sci U S A. 2002;99:11375–11380.

33. Lin T, Gu J, Huang C, Zheng S, Lin X, Xie L, Lin D. (1)H NMR-Based Analysis of Serum Metabolites in Monocrotaline-Induced Pulmonary Arterial Hypertensive Rats. Dis Markers. 2016;2016:5803031.

